# Sound of Aging: Large-Scale Evidence for a Voice-Based Biological Clock

**DOI:** 10.64898/2026.04.05.26350190

**Authors:** David Krongauz, Yanir Marmor, Arad Zulti, Anastasia Godneva, Adina Weinberger, Eran Segal

**Author notes:** Correspondence (D.K.) (E.S.). These authors contributed equally to this work (David Krongauz, Yanir Marmor).

## Abstract

Using 30-second voice recordings from 7,081 adults aged 40–70, we trained gender-specific models to estimate voice-predicted age (Voice Age). Voice Age correlated with chronological age comparably to established omic and physiological aging clocks, while capturing an independent dimension of biological aging. Accelerated vocal aging showed association with higher adiposity, impaired sleep physiology, and cardiometabolic risk markers, supporting voice as a scalable, non-invasive functional aging biomarker.

## Main

Aging manifests heterogeneously across molecular, physiological, and functional domains, such that chronological age is a poor proxy for individual aging trajectories ^1–4^. Growing evidence indicates that aging proceeds asynchronously across biological systems; molecular damage does not always translate linearly into organism-level functional decline^1,3,5^. Biological aging clocks derived from epigenetic, proteomic, and imaging data have improved risk stratification, yet many remain costly, invasive, and difficult to deploy at scale^3,6,7^. While functional measures like gait speed and grip strength predict mortality independently of molecular age, they are difficult to standardize and require in-person assessment^8–10^. Therefore, there is a need for scalable functional biomarkers that can bridge the gap between cellular damage and phenotypic decline.

The human voice offers a promising candidate to address this gap^11,12^. Speech production is a complex motor task requiring millisecond-scale coordination of respiratory pressure, laryngeal tension, and articulatory kinematics, systems that undergo well-characterized structural and functional changes with age. Age-related sarcopenia, alterations in connective-tissue elasticity, and subtle neuromotor decline collectively shape acoustic features such as spectral stability and temporal variability^13,14^. We therefore hypothesized that the human voice, a ubiquitous and low-cost biosignal, encodes a measurable signature of functional biological aging that is partially independent of molecular and imaging-based clocks.

To test this, we analyzed 10,434 standardized 30-second voice recordings from 8,800 adults aged 40–70 years enrolled in a population-based cohort that combines broad demographic representation with extensive multi-omic, clinical, and physiological phenotyping^15^. Following strict quality control to remove technical artifacts and corrupt recordings, the final analytic dataset comprised 7,784 recordings from 7,081 individuals (3,400 females and 3,681 males). Using gender-stratified models trained on deep speech embeddings, we defined Voice Age (VA) as the age predicted from vocal features alone, and Voice Age acceleration (ΔVA) as the deviation of VA from chronological age, reflecting whether an individual’s voice appears biologically older or younger than expected^16^.

VA showed strong concordance with chronological age. In both genders, VA correlated with chronological age (Pearson r ≈ 0.75 in females and r ≈ 0.67 in males), explaining 56 ± 0.22% and 45 ± 0.17% of age variance in cross-validation, respectively. This yielded mean absolute errors (MAEs) of 3.89 ± 0.01 and 4.35 ± 0.01 years (see Fig. 1). This performance is comparable to that of established omics and physiological aging clocks that were evaluated on the same cohort using similar cross-validation procedures^4^. Models trained on deep speech embeddings outperformed classical acoustic feature sets, including mel-frequency cepstral coefficients and perturbation measures (jitter and shimmer), indicating that higher-order temporal and spectral structure contribute meaningfully to vocal age estimation (see Supplementary Methods S5 and Fig. S1).

**Fig. 1.**
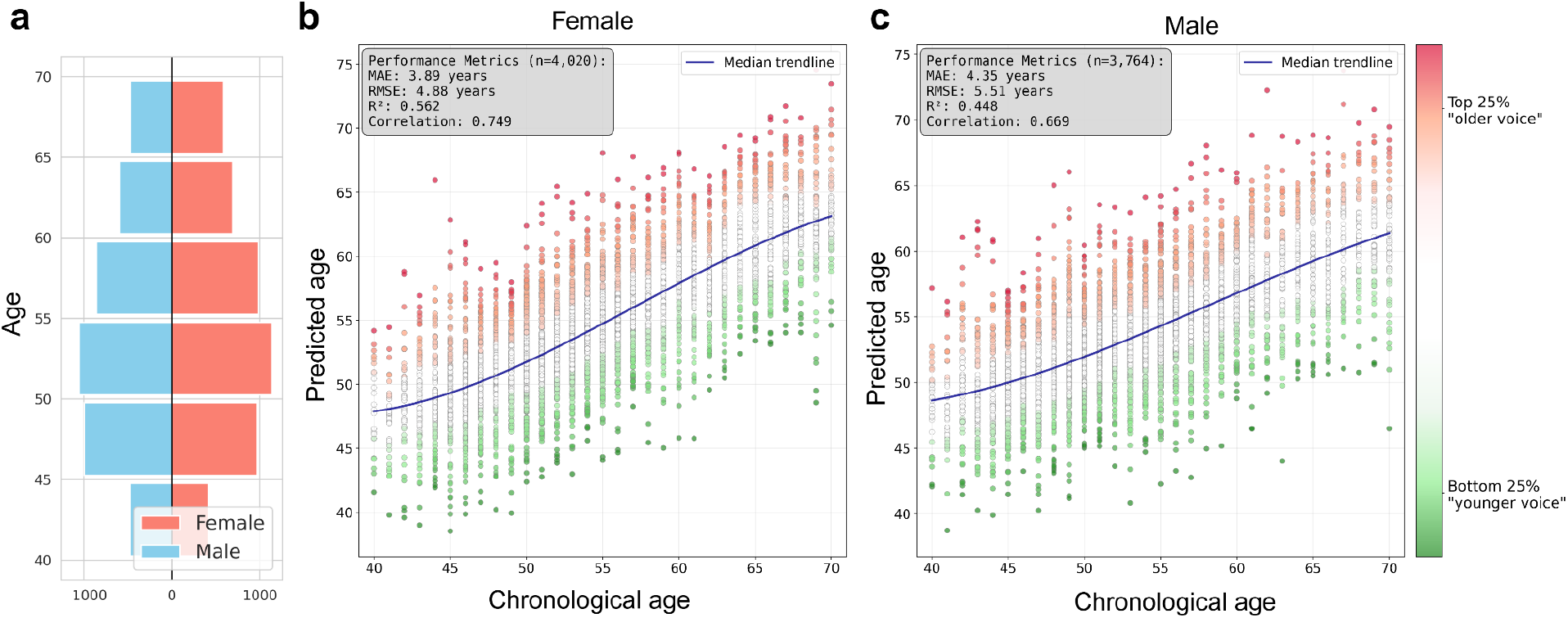
Voice Age prediction performance and cohort distribution. **a**, Population pyramid displaying the age and gender distribution of the analytic cohort (n = 7,081 participants). **b,c**, Scatter plots illustrating the concordance between predicted Voice Age and chronological age for females (**b**; n = 4,020 recordings) and males (**c**; n = 3,764 recordings). Performance metrics for females were Pearson r = 0.75, R^2^ = 0.56, mean absolute error (MAE) = 3.89 years, and root mean squared error (RMSE) = 4.88 years. Metrics for males were Pearson r = 0.67, R^2^ = 0.45, MAE = 4.35 years, and RMSE = 5.51 years. The blue solid line represents the median predicted age trend. Data points are colored according to Voice Age acceleration (ΔVA) quartiles: red indicates the top 25% (accelerated or ‘older-sounding’ aging), green indicates the bottom 25% (decelerated or ‘younger-sounding’ aging).

To evaluate the biological relevance of ΔVA, we investigated whether individuals with accelerated or decelerated vocal aging exhibited differences across multiple physiological systems. Within each two-year age bin and separately by gender, participants in the top and bottom ΔVA quartiles were compared (Mann–Whitney tests) across clinical, biochemical, imaging, and lifestyle phenotypes (Fig. 2). This analysis showed consistent associations across multiple phenotypic domains, supporting ΔVA as a marker of systemic aging-related variation beyond vocal tract features alone.

**Fig. 2.**
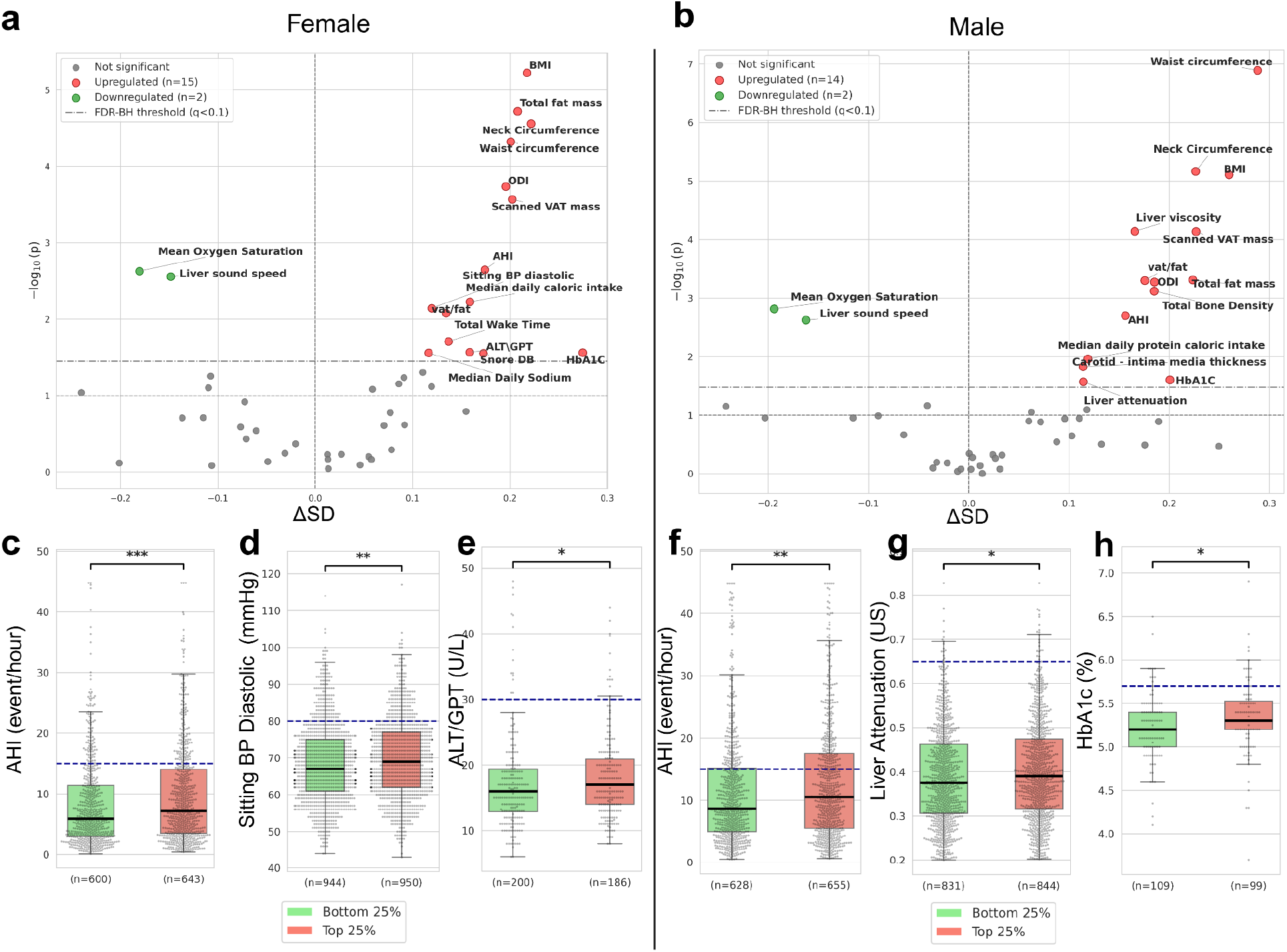
Physiological associations of accelerated vocal aging. **a,b**, Volcano plots displaying phenome-wide differences between individuals with accelerated versus decelerated vocal aging in females (**a**) and males (**b**). The x-axis represents the standardized mean difference (ΔSD) between the top (accelerated) and bottom (decelerated) Voice Age acceleration (ΔVA) quartiles. The y-axis shows the negative log10-transformed P value (two-sided Mann–Whitney U test). Features significantly elevated in accelerated agers (FDR q < 0.1) are colored red; features significantly lower are colored green. **c**–**h**, Box plots comparing representative biomarkers between the bottom 25% (green, ‘younger-sounding’) and top 25% (red, ‘older-sounding’) ΔVA quartiles. Horizontal dashed blue lines indicate established clinical thresholds for abnormality: AHI > 15 events/h (obstructive sleep apnea),^17^ diastolic blood pressure > 80 mmHg (hypertension),^18^ ALT > 30 U/L (possible liver pathology in females),^19^ liver attenuation > 0.65 dB/cm/MHz (hepatic steatosis),^20^ and HbA1c > 5.7% (prediabetes).^21^ Box centers represent the median, bounds indicate the interquartile range (IQR), and whiskers extend to 1.5 × IQR. Significance levels: *P < 0.05, **P < 0.01, ***P < 0.001.

In both genders, accelerated vocal aging was associated with higher adiposity across multiple measures, including body mass index, waist and neck circumference, total and visceral fat mass, and liver fat indices (Fig. 2). Markers of impaired sleep physiology were also consistently elevated, including higher apnea–hypopnea and oxygen-desaturation indices, increased snoring intensity, greater total nocturnal wake time, and lower mean nocturnal oxygen saturation.

Additional associations extended beyond adiposity and sleep. In men, accelerated vocal aging was associated with increased carotid intima–media thickness, elevated white blood cell count, altered liver ultrasound properties (reduced sound speed and increased attenuation), and higher HbA1c levels. In women, associations included higher diastolic blood pressure, elevated liver enzymes (ALT), greater caloric intake, and adverse glycemic markers. While several effects were modest in magnitude, their directionality was consistent across related phenotypes and physiological systems (Supplementary Table S1).

Conversely, individuals with younger-sounding voices exhibited more favorable profiles across these domains, including lower adiposity, preserved sleep oxygenation, and healthier liver imaging characteristics. Together, these findings indicate that ΔVA captures coordinated variation across cardiometabolic, sleep, vascular, and inflammatory systems, supporting its relevance as a marker of functional biological aging.

A central question in geroscience is whether new aging clocks capture information distinct from existing measures^5,7^. We compared Voice Age with eight clocks constructed from metabolomic, imaging, physiological, lifestyle, and microbiome data, all evaluated under an identical cross-validation framework. In terms of predictive power, Voice Age emerged as a highly robust biomarker. It ranked as the second-best single-modality predictor in both genders (R^2^ ≈ 0.56 in females, 0.45 in males), outperforming established physiological and imaging-based clocks, including sleep, body composition (DEXA), and retinal imaging, and trailing only serum metabolomics (Fig. 3a). Despite this high predictive accuracy, Voice Age shared only partial overlap with other modalities (Pearson r range: 0.16–0.58), suggesting it captures a unique dimension of the aging process. Interestingly, Voice Age showed the strongest alignment with MS metabolomics (r = 0.58 females; 0.43 males) and sleep-based clocks (r = 0.55 females; 0.43 males) (see Fig. 3, Supplementary Fig. S3). Conversely, weaker correlations were observed with standard blood biochemistry and microbiome profiles, indicating that vocal aging is largely independent of these specific biological compartments. To test for complementarity, we evaluated multimodal models combining voice features with each non-acoustic modality. The addition of voice consistently improved predictive performance (positive synergy) for seven out of eight modalities, most notably improving diet and sleep-based clocks, with the sole exception of MS metabolomics, where no additional gain was observed (Supplementary Fig. S2).

**Fig. 3.**
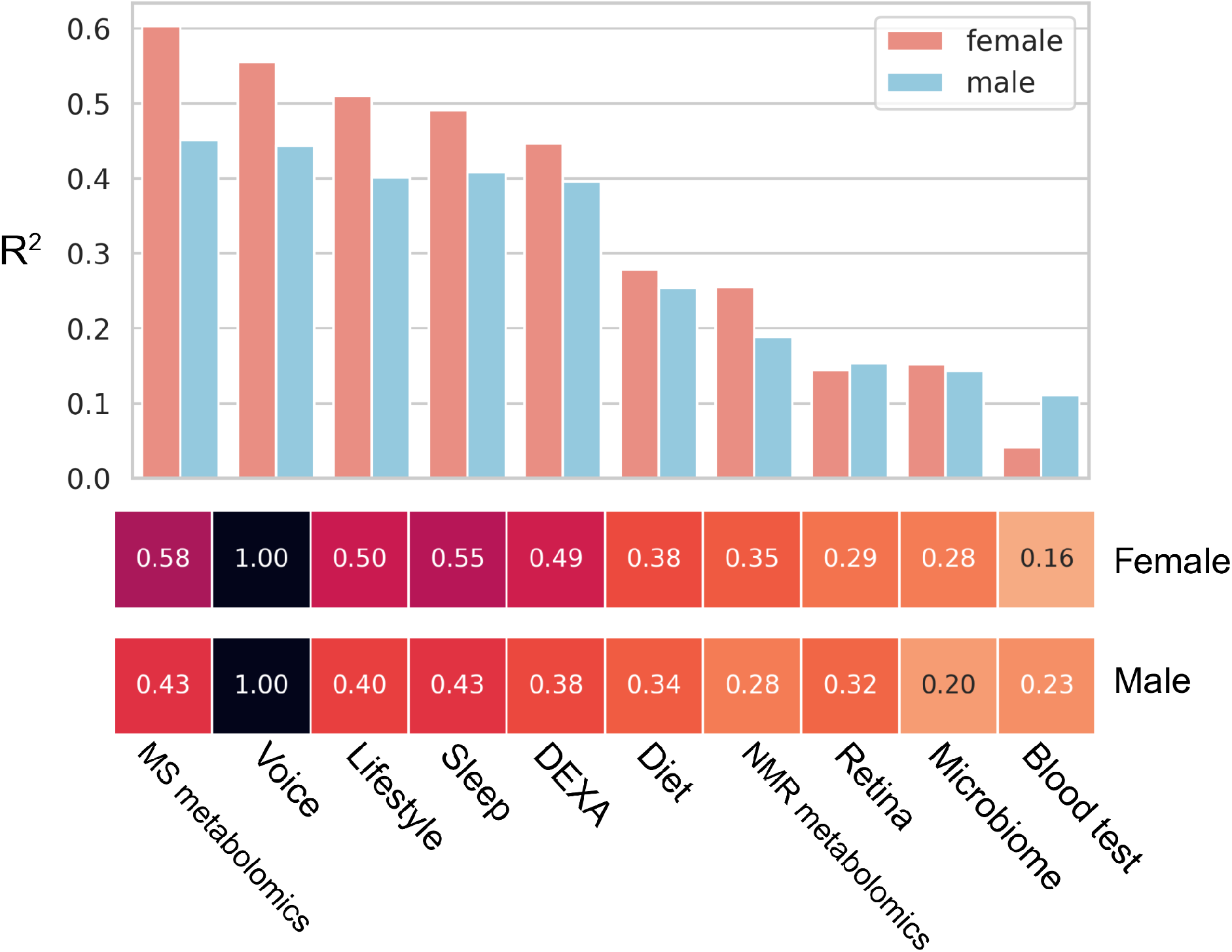
Benchmarking Voice Age against multi-modal biological aging clocks. Bar chart comparing the predictive performance (mean R^2^, 5-fold CV) of the voice-based model against eight other biological age estimators trained on the same cohort. Models are ranked by performance in females. Voice outperforms imaging (DEXA, Retina), physiological (Sleep), and lifestyle-based clocks, performing second only to metabolomics. Parentheses indicate the type of model that was best performing for each specific modality. Below, the heatmaps display the pairwise Pearson correlation coefficients (r) between predicted Voice Age and the other biological ages.

This study has limitations. First, the cross-sectional design precludes causal inference; longitudinal data will be required to determine whether within-person changes in Voice Age track aging or respond to interventions. Second, recordings were collected under standardized clinical conditions in a single cohort, and generalizability across languages, recording environments, and devices remains to be established. Third, although deep speech embeddings enable robust age estimation, their latent representations are only partially interpretable; linking specific acoustic dimensions to underlying physiology will require targeted experimental studies.

Despite these limitations, our findings show that a brief voice sample encodes aging-related information that complements established molecular and physiological clocks.^11,12^ Because speech can be collected non-invasively and repeatedly at low cost, voice-based measures offer a practical route to repeated, population-scale assessment of functional aging. Integrating acoustic measures with molecular and imaging data may enable more comprehensive characterization of aging trajectories and physiological resilience in research and clinical settings.

## Methods

### 1. Study design and participants

This study was conducted within the Human Phenotype Project (HPP), an ongoing cohort study designed to characterize biological and functional aging through extensive clinical, molecular, and physiological phenotyping. Participants were recruited between 2019 and 2025 at a single clinical research site in Israel and completed standardized baseline assessments during scheduled clinic visits.

Eligible participants were Hebrew-speaking adults aged 40–70 years with a valid 30-second voice recording and reported chronological age. No additional exclusion criteria were applied beyond voice-recording quality control and the availability of age and gender. After quality control (see Fig. S4), the final analytic sample comprised 7,081 individuals (3,681 females and 3,400 males).

All participants provided written informed consent, and the study was approved by the Institutional Review Board of the Weizmann Institute of Science (approval no. 2392-4), in accordance with the Declaration of Helsinki.

### 2. Voice data acquisition and preprocessing

Voice recordings were collected during standardized clinic visits as part of the HPP assessment protocol. Participants performed a single 30-second fluent counting task (counting aloud from 1 to 30 at a comfortable pace) while seated in a quiet clinical examination room.

Recordings were obtained under controlled clinical conditions using a fixed protocol and were downsampled to 16 kHz, single-channel (mono) prior to analysis. Leading and trailing silence was removed using an automated energy-based procedure, and peak normalization was applied on a per-recording basis.

An automated quality-control pipeline was applied to identify recordings affected by technical artifacts or incomplete task performance. Of 10,434 recordings collected, 7,784 recordings from 7,081 participants passed quality control and were retained for downstream analyses. Participant flow and exclusion counts are summarized in Supplementary Fig. S4. Additional details are provided in Supplementary Methods S1–S2.

### 3. Acoustic feature extraction

Acoustic representations were derived using WavLM-Large, a self-supervised speech representation model trained on large-scale multilingual audio corpora. For each 30-second recording, the pre-trained encoder was applied without task-specific adaptation or parameter updates.

The encoder produced frame-level embeddings with a dimensionality of 1024, which were aggregated across the temporal dimension using mean pooling to obtain a single, fixed-length representation per recording. For benchmarking purposes, a set of classical acoustic features, including mel-frequency cepstral coefficients (MFCCs) and perturbation-based measures such as jitter and shimmer, was also extracted but not used in the primary Voice Age model. Additional implementation details are provided in Supplementary Methods S3.

### 4. Voice Age model training

Voice Age prediction models were trained separately for females and males to account for known gender-specific differences in vocal anatomy and aging trajectories. Chronological age was predicted from high-dimensional acoustic embeddings using ridge regression.

Model performance was evaluated using five-fold cross-validation, with folds stratified to preserve the age distribution within each gender and grouped by participant ID, in case of multiple visits per participant. For each fold, models were trained on 80% of the data and evaluated on the held-out 20%. Performance was assessed using the Pearson correlation coefficient (r), coefficient of determination (R^2^), mean absolute error (MAE), and root mean squared error (RMSE). Reported metrics represent averages across cross-validation folds.

For each participant, Voice Age (VA) was defined as the age predicted by the corresponding gender-specific model. Voice Age acceleration (ΔVA) was computed as ΔVA = VA − CA, where CA denotes chronological age. No additional residualization or post hoc age-bias correction was applied.

### 5. Phenotype definitions

Phenotypic measures were derived from standardized assessments conducted as part of the HPP protocol. Analyses focused on predefined classes of physiological and clinical phenotypes relevant to aging, including anthropometrics, sleep physiology, cardiometabolic markers, and imaging-derived measures.

Anthropometric phenotypes included body mass index, waist and neck circumference, and body composition indices. Sleep-related phenotypes encompassed measures of sleep-disordered breathing, nocturnal oxygenation, and sleep continuity. Cardiometabolic markers included blood-based indicators of metabolic and inflammatory status, as well as vascular measurements. Imaging-derived phenotypes included ultrasound- and densitometry-based assessments of organ structure and body composition.

Detailed descriptions of measurement protocols, device specifications, and quality-control procedures are provided in the primary Human Phenotype Project cohort description and in Supplementary Methods ^15^

### 6. Comparison of aging clocks

To assess non-redundant aging-related information, Voice Age was compared with a panel of eight biological-age clocks derived from non-acoustic data modalities available within the HPP, including metabolomic, imaging, physiological, lifestyle, dietary, microbiome, and sleep-derived features.

All comparison clocks were trained de novo within the same cohort using the same modeling pipeline and cross-validation framework as the voice-based model. Predicted ages were expressed on the chronological-age scale to facilitate correlation analyses and multimodal integration.

Cross-clock relationships were assessed using pairwise correlations between predicted ages. Complementarity was evaluated by training multimodal models combining voice-derived features with each non-acoustic modality and quantifying changes in predictive performance relative to single-modality clocks. Additional details are provided in Supplementary Methods S4.

### 7. Statistical analysis

Associations between Voice Age, chronological age, and other aging clocks were evaluated using Pearson correlation coefficients, with analyses conducted separately by gender unless otherwise stated.

To assess biological differences associated with variation in vocal aging, participants were stratified within two-year chronological age bins and by gender. Within each bin, individuals were ranked by Voice Age acceleration (ΔVA) and assigned to quartiles. Participants in the top quartile were compared with those in the bottom quartile.

Group differences across phenotypes were evaluated using two-sided Mann–Whitney U tests. Multiple hypothesis testing was controlled using the Benjamini–Hochberg false discovery rate procedure.

## Data Availability

All data produced in the present study are available to qualified researchers through the Human Phenotype Project via a formal application process administered by the HPP data access committee.

https://www.pheno.ai/

## 8. Data and code availability

The data used in this study are available to qualified researchers through the Human Phenotype Project via a formal application process administered by the HPP data access committee. Code used for model training and analysis is available via GitHub at https://github.com/kronga/Sound_of_Aging

## 9. Acknowledgements

We thank the members of the Segal group for fruitful discussions. ES is supported by the Crown Human Genome Center; the Larson Charitable Foundation New Scientist Fund; the Else Kröner Fresenius Foundation; the White Rose International Foundation; the Ben B. and Joyce E. Eisenberg Foundation; the Nissenbaum Family; Marcos Pinheiro de Andrade and Vanessa Buchheim; Lady Michelle Michels; Aliza Moussaieff; and grants funded by the Minerva Foundation, with funding from the Federal German Ministry for Education and Research and by the European Research Council and the Israel Science Foundation. Views and opinions expressed are, however, those of the author(s) only and do not necessarily reflect those of the European Union. Neither the European Union nor the granting authority can be held responsible for them. The funders had no role in study design, data collection and analysis, decision to publish or preparation of the manuscript.

## 10. Author contributions

D.K. and Y.M. conceived the project, designed and conducted all analyses, interpreted the results, and wrote the paper. A.Z. and A.G. conducted analyses and interpreted results. A.W. interpreted results and supervised data acquisition. E.S. conceived and directed the project, designed the analyses, interpreted the results, and wrote the paper. All authors read and approved the final manuscript.

## References

1. López-Otín, C., Blasco, M. A., Partridge, L., Serrano, M. & Kroemer, G. Hallmarks of aging: An expanding universe. Cell 186, 243–278 (2023).

2. Jylhävä, J., Pedersen, N. L. & Hägg, S. Biological Age Predictors. eBioMedicine 21, 29–36 (2017).

3. Rutledge, J., Oh, H. & Wyss-Coray, T. Measuring biological age using omics data. Nat. Rev Genet. 23, 715–727 (2022).

4. Reicher, L. et al. Phenome-wide associations of human aging uncover sex-specific dynamics. Nat. Aging 4, 1643–1655 (2024).

5. Drewelies, J. et al. There Are Multiple Clocks That Time Us: Cross-Sectional and Longitudinal Associations Among 14 Alternative Indicators of Age and Aging. J. Gerontol. Ser. A 80, glae244 (2025).

6. Lu, A. T. et al. DNA methylation GrimAge strongly predicts lifespan and healthspan. Aging 11, 303–327 (2019).

7. Min, M., Egli, C., Dulai, A. S. & Sivamani, R. K. Critical review of aging clocks and factors that may influence the pace of aging. Front. Aging 5, (2024).

8. Petersen, C. L., Christensen, B. C. & Batsis, J. A. Weight management intervention identifies association of decreased DNA methylation age with improved functional age measures in older adults with obesity. Clin. Epigenetics 13, 46 (2021).

9. Turimov Mustapoevich, D. & Kim, W. Machine Learning Applications in Sarcopenia Detection and Management: A Comprehensive Survey. Healthcare 11, 2483 (2023).

10. Goldman, N., Glei, D. A., Rosero-Bixby, L.Chiou, S.-T. & Weinstein, M. Performance-based measures of physical function as mortality predictors: Incremental value beyond self-reports. Demogr. Res. 30, 227–252 (2014).

11. Härmä, A. et al. Survey on biomarkers in human vocalizations. Preprint at 10.48550/arXiv.2407.17505 (2024).

12. Fagherazzi, G., Fischer, A., Ismael, M. & Despotovic, V. Voice for Health: The Use of Vocal Biomarkers from Research to Clinical Practice. Digit. Biomark. 5, 78–88 (2021).

13. Dehqan, A., Scherer, R. C., Dashti, G., Ansari-Moghaddam, A. & Fanaie, S. The Effects of Aging on Acoustic Parameters of Voice. Folia Phoniatr. Logop. 64, 265–270 (2013).

14. Rojas, S., Kefalianos, E. & Vogel, A. How Does Our Voice Change as We Age? A Systematic Review and Meta-Analysis of Acoustic and Perceptual Voice Data From Healthy Adults Over 50 Years of Age. J. Speech Lang. Hear. Res. 63, 533–551 (2020).

15. Reicher, L. et al. Deep phenotyping of health–disease continuum in the Human Phenotype Project. Nat. Med. 31, 3191–3203 (2025).

16. Chen, S. et al. WavLM: Large-Scale Self-Supervised Pre-Training for Full Stack Speech Processing. IEEE J. Sel. Top. Signal Process. 16, 1505–1518 (2022).

17. Sateia, M. J. International Classification of Sleep Disorders-Third Edition. Chest 146, 1387–1394 (2014).

18. 2017 Guideline for High Blood Pressure in Adults. American College of Cardiology https://www.acc.org/latest-in-cardiology/ten-points-to-remember/2017/11/09/11/41/ http://www.acc.org%2flatest-in-cardiology%2ften-points-to-remember%2f2017%2f11%2f09%2f11%2f41%2f2017-guideline-for-high-blood-pressure-in-adults.

19. Valenti, L. et al. Definition of Healthy Ranges for Alanine Aminotransferase Levels: A 2021 Update. Hepatol. Commun. 5, 1824 (2021).

20. Dioguardi Burgio, M. et al. Quantification of hepatic steatosis with ultrasound: promising role of attenuation imaging coefficient in a biopsy-proven cohort. Eur. Radiol. 30, 2293–2301 (2020).

21. 2. Diagnosis and Classification of Diabetes: Standards of Care in Diabetes—2024. Diabetes Care 47, S20–S42 (2023).

