## Supplementary Materials for "Sound of Aging: Large-Scale Evidence for a Voice-Based Biological Clock"

### Supplementary material - Sound of Aging

##### Supplementary Methods S1 – Voice recording protocol

Voice recordings were collected during standardized clinic visits as part of the Human Phenotype Project. Participants performed a single 30-second fluent counting task while seated in a quiet clinical examination room. A trained research staff member was present during recording to ensure task compliance and consistency across sessions.

Audio was captured using a fixed microphone stand placed on a desk in front of the participant, resulting in a consistent mouth–microphone distance across recordings. Recordings were initially acquired at high resolution and subsequently downsampled to 16 kHz, single-channel (mono) prior to downstream analysis. No additional speech tasks were administered.

##### Supplementary Methods S2 – Voice quality control classifier

Automated quality control was applied to identify recordings affected by technical artifacts. A supervised Random Forest classifier was trained on a manually annotated subset of 488 recordings. Annotations were provided by a single expert annotator and labeled recordings as either technically acceptable or faulty based on the presence of clipping, excessive background noise, signal dropouts, or incomplete task execution.

Mel-frequency cepstral coefficients (MFCCs) were extracted from each recording and used as input features. The annotated dataset comprised approximately 70% high-quality recordings and 30% faulty recordings. Classifier performance was evaluated using five-fold cross-validation, yielding a mean area under the receiver operating characteristic curve (AUC) of $0.95 \pm 0.04$. Recordings with a predicted probability greater than 0.5 of being faulty were excluded from downstream analyses.

##### Supplementary Methods S3 – Acoustic embeddings

Acoustic embeddings were extracted using the WavLM-Large self-supervised speech representation model. The pre-trained encoder was applied using default settings without task-specific adaptation or fine-tuning. Embeddings were extracted exclusively from the final transformer layer.

For each recording, frame-level embeddings (with a dimensionality of 1024) were aggregated across time using simple mean pooling to obtain a single, fixed-length embedding. No additional padding or truncation beyond silence trimming was applied prior to embedding extraction.

**Supplementary Methods S4 – Comparison of aging clocks**

A panel of eight biological-age clocks was constructed from non-acoustic data modalities available within the Human Phenotype Project, including metabolomic, imaging, physiological, lifestyle, dietary, microbiome, and sleep-derived feature sets.

All comparison clocks were trained de novo within the same cohort using identical cross-validation fold assignments as the voice-based model. Chronological age was predicted separately for each modality using either ridge regression or LightGBM models, depending on modality characteristics. Hyperparameters were fixed a priori and not tuned within cross-validation folds. No additional feature normalization or missing-value imputation was applied.

All clocks were trained in a gender-stratified manner. Predicted ages were expressed on the chronological-age scale to enable direct comparison, correlation analysis, and multimodal integration with Voice Age.

**Supplementary Methods S5 - Comparison of acoustic feature representations.**

To assess the contribution of different speech feature sets, we trained parallel age-prediction models using (i) 13 Mel-frequency cepstral coefficients (MFCCs), (ii) a standard set of 88 hand-crafted acoustic features extracted with the openSMILE library,[1] and (iii) deep self-supervised speech embeddings from WavLM-Large (1024 dimensions).[2] All feature sequences were temporally pooled by taking the mean across time before model training. As shown in Supplementary Fig. S1, models based on WavLM embeddings substantially outperformed both MFCC and openSMILE-based representations, indicating that high-level learned speech representations capture age-related vocal information more effectively than classical acoustic features.

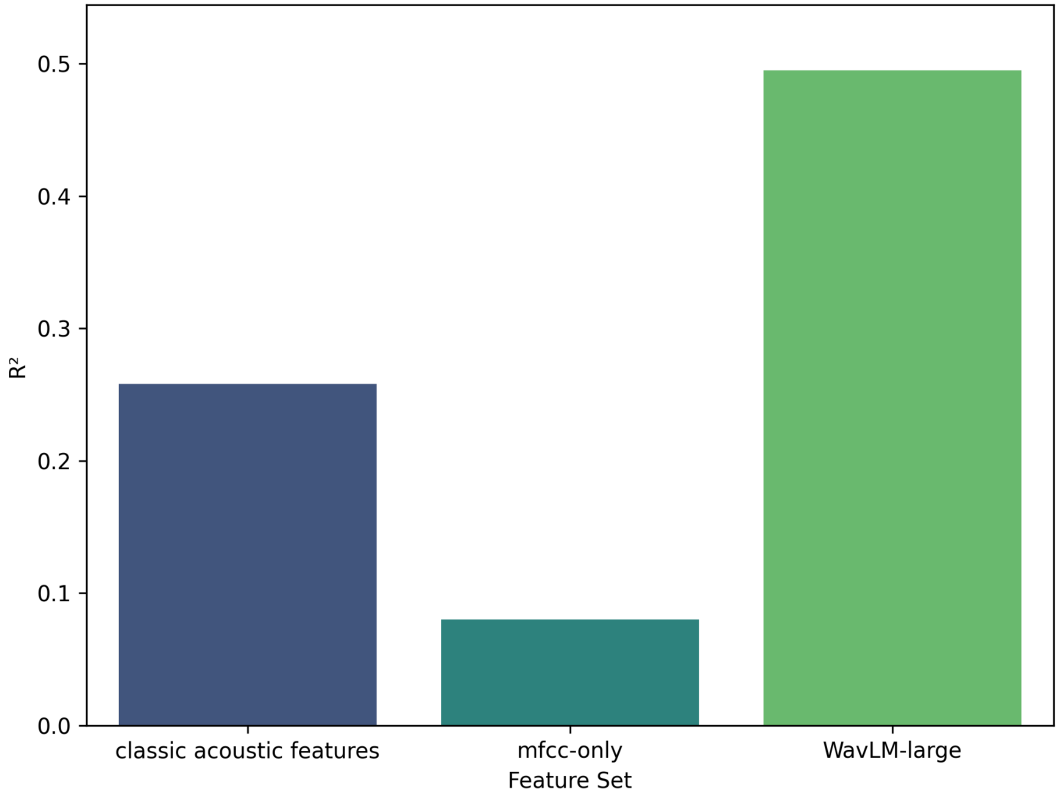


**Fig. S1 | Comparison of feature representations for voice-based age prediction.**

Predictive performance (cross-validated R^2) of models trained on different speech feature sets, with all features temporally pooled by the mean. Classical acoustic features derived from the openSMILE toolkit (88 features) and MFCC-only features (13 coefficients) show limited explanatory power, whereas deep speech embeddings from WavLM-large (1024 dimensions) substantially improve age prediction performance.

**S2**

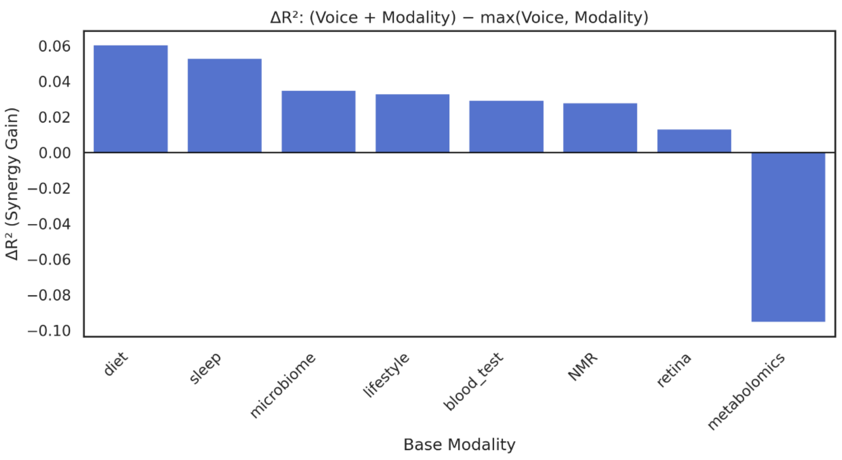


**S3a**

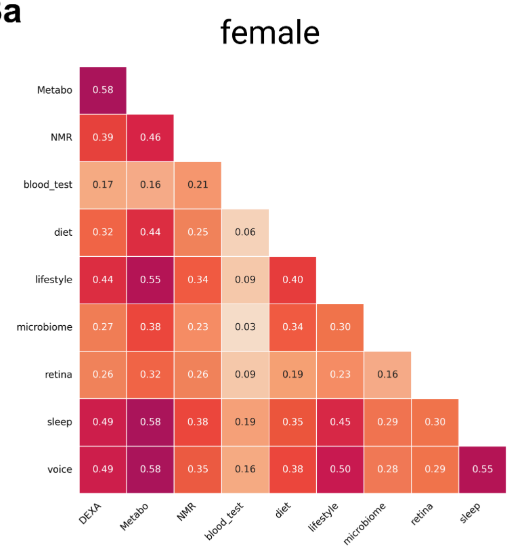


**S3b**

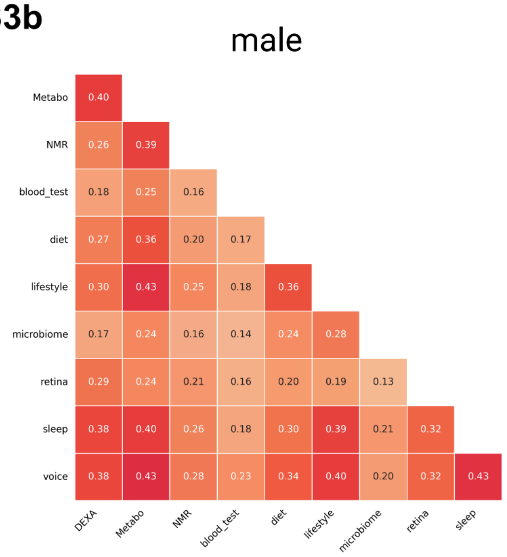


**Figure S2 | Complementarity of Voice Age with other aging clocks.**

Synergy gain ($\Delta R^2$) obtained by adding voice features to models based on each non-acoustic aging modality, computed as the improvement in cross-validated $R^2$ of the combined model (voice + modality) over the better of the two single-modality models. Positive values indicate that voice contributes complementary information beyond the original clock. Voice features improved predictive performance for seven of eight modalities, with the largest gains observed for diet- and sleep-based clocks, indicating a substantial non-redundant aging signal. No gain was observed for MS-based metabolomics.

**Supplementary Figure S3 | Cross-modal correlations between Voice Age and other aging clocks.**

a, Female; b, Male. Pairwise Pearson correlation coefficients between Voice Age and eight aging clocks derived from metabolomic, imaging, physiological, lifestyle, and microbiome data, evaluated under an identical cross-validation framework. In both sexes, Voice Age shows moderate correlations with other modalities, indicating partial overlap alongside substantial independence. The strongest associations are observed with MS-based metabolomics and sleep-derived clocks, whereas weaker correlations with blood biochemistry and microbiome-based clocks suggest that vocal aging captures aging-related information largely distinct from these biological compartments.

**S4**

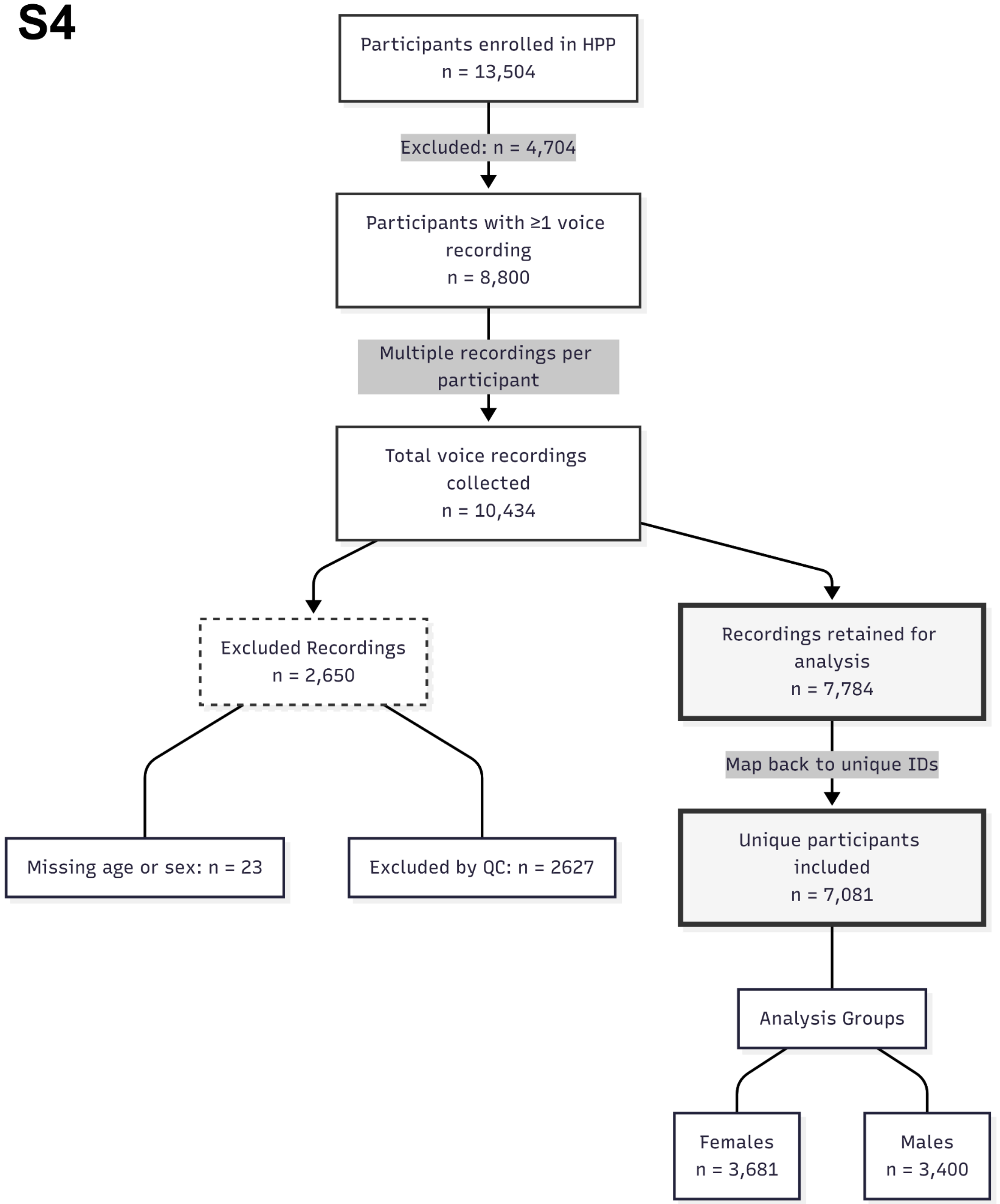


**Supplementary Figure S4 | Participant flow and construction of the voice analysis cohort.**

Flow diagram describing participant inclusion and exclusion from the Human Phenotype Project (HPP) voice dataset. Of 13,504 enrolled participants, 8,800 had at least one voice recording, yielding 10,434 total recordings. After excluding recordings with missing age or sex information and those that failed voice quality control, 7,784 recordings were retained and mapped back to unique participant identifiers. The final analytic cohort comprised 7,081 individuals (3,681 females and 3,400 males) aged 40–70 years, who were included in all subsequent analyses.

| Counts | | n=3,400 | | n=3,681 | |
|---|---|---|---|---|---|
| **Feature** | **Unit** | **Male Mean** | **Male Std** | **Female Mean** | **Female Std** |
| Age | Years | 53.9 | 7.45 | 54.7 | 7.38 |
| **Smoking status** | | | | | |
| Current smokers | % | 10.06 | | 10.19 | |
| Former smokers | % | 28.43 | | 29.05 | |
| **Vitals & Anthropometrics** | | | | | |
| Waist circumference | cm | 93.39 | 10.73 | 82.33 | 10.94 |
| Sitting BP systolic | mmHg | 124.7 | 15.22 | 114.28 | 16.73 |
| Sitting BP diastolic | mmHg | 73.39 | 10.53 | 69.19 | 10.89 |
| Neck Circumference | cm | 38.63 | 2.98 | 32.79 | 2.54 |
| BMI | $kg/m^2$ | 26.66 | 3.8 | 25.73 | 4.43 |
| **Blood Test (BT)** | | | | | |
| Platelet count (BT) | $10^3/\mu L$ | 226.65 | 50.09 | 257.2 | 56.65 |
| HDL cholesterol (BT) | mg/dL | 51.48 | 11.36 | 62.84 | 13.84 |
| HbA1C (BT) | % | 5.31 | 0.44 | 5.34 | 0.38 |
| WBC (BT) | $10^3/\mu L$ | 6.24 | 1.43 | 6.19 | 1.51 |
| Total cholesterol (BT) | mg/dL | 187.36 | 37.45 | 198.25 | 35.94 |
| ALT\GPT (BT) | U/L | 24.72 | 9.97 | 17.93 | 7.44 |
| LDL cholesterol (BT) | mg/dL | 118.73 | 29.49 | 116.58 | 30.64 |
| AST\GOT (BT) | U/L | 25.54 | 6.56 | 21.85 | 5.88 |
| Albumin (BT) | g/dL | 4.38 | 0.24 | 4.22 | 0.23 |
| GGT (BT) | U/L | 26.05 | 13.97 | 18.47 | 11.61 |
| CRP (BT) | mg/dL | 0.51 | 0.87 | 0.42 | 0.63 |
| Triglycerides (BT) | mg/dL | 109.94 | 56.08 | 97.63 | 48.1 |
| eGFR (BT) | $mL/min/1.73 m^2$ | 83.09 | 13.79 | 82.84 | 14.2 |

| | | | | | |
|---|---|---|---|---|---|
| **Sleep Monitor (SM)** | | | | | |
| SleepEfficiancy | % | 87.59 | 5.13 | 88.73 | 4.51 |
| Total Sleep Time (SM) | Hours | 5.82 | 1.19 | 6.1 | 1.19 |
| Snore DB | dB | 40.84 | 0.89 | 40.68 | 0.82 |
| RDI (SM) | Events/h | 18.75 | 10.02 | 13.53 | 8.09 |
| Rem Latency | Seconds | 4865.71 | 2254.62 | 4735.33 | 2083.14 |
| ODI (SM) | Events/h | 4.94 | 4.84 | 3.35 | 3.75 |
| AHI (SM) | Events/h | 12.68 | 9.75 | 8.92 | 7.93 |
| Total Wake Time (SM) | Hours | 0.8 | 0.34 | 0.76 | 0.31 |
| Mean Oxygen Saturation (SM) | % | 94.61 | 1.1 | 94.8 | 1.14 |
| **Body Composition (DXA)** | | | | | |
| Android tissue fat percent (DXA) | % | 12.78 | 17.49 | 15.19 | 19.93 |
| Total Bone Density | g/cm² | 1.27 | 0.12 | 1.13 | 0.11 |
| Total fat mass (DXA) | g | 22862.6 | 8080.3 | 25563.7 | 8533.6 |
| Scanned VAT mass (DXA) | g | 1167.77 | 743.5 | 647.71 | 456.54 |
| vat/fat | Ratio | 0.047 | 0.021 | 0.023 | 0.012 |
| **Ultrasound (US)** | | | | | |
| Liver viscosity (US) | Pa·s | 1.67 | 0.21 | 1.56 | 0.21 |
| Liver elasticity (US) | kPa | 5.27 | 1.04 | 4.51 | 0.82 |
| Liver attenuation (US) | dB/cm/MHz | 0.4 | 0.12 | 0.4 | 0.12 |
| Carotid - intima media thickness (US) | mm | 0.62 | 0.12 | 0.6 | 0.1 |
| Liver sound speed (US) | m/s | 1565.94 | 37.43 | 1552.56 | 36.72 |
| **Diet Log (DL)** | | | | | |
| Median daily caloric intake (DL) | kcal | 1682.08 | 507.72 | 1411.69 | 438.14 |
| Median daily carbohydrate caloric intake (DL) | Ratio | 0.41 | 0.1 | 0.43 | 0.1 |
| Median daily lipid caloric intake (DL) | Ratio | 0.39 | 0.08 | 0.39 | 0.08 |
| Median daily protein caloric intake (DL) | Ratio | 0.19 | 0.05 | 0.18 | 0.04 |
| Median Daily Sodium (DL) | g | 1.53 | 0.38 | 1.48 | 0.38 |
| Daily Fiber (DL) | kg | 0.011 | 0.004 | 0.012 | 0.005 |
| Daily Folate (DL) | mg | 0.189 | 0.065 | 0.195 | 0.063 |
| **Fundus Image (FI)** | | | | | |
| Average width (FI) | nm | 16832.1 | 1067.9 | 17351.7 | 1196.8 |

| | | | | | |
|---|---|---|---|---|---|
| Fractal dimension (FI) | Dimensionless | 1.53 | 0.02 | 1.52 | 0.02 |
| Vessel density (FI) | Ratio | 0.075 | 0.007 | 0.076 | 0.007 |

**Supplementary Table S1 | Baseline characteristics of the study cohort.** Demographic, clinical, and physiological characteristics of the 7,081 participants included in the final analysis, stratified by gender.

#### References


1. Eyben, F., Wöllmer, M. & Schuller, B. Opensmile: the munich versatile and fast open-source audio feature extractor. in *Proceedings of the 18th ACM international conference on Multimedia* 1459–1462 (Association for Computing Machinery, New York, NY, USA, 2010). doi:10.1145/1873951.1874246.
2. Chen, S. *et al.* WavLM: Large-Scale Self-Supervised Pre-Training for Full Stack Speech Processing. *IEEE J. Sel. Top. Signal Process.* **16**, 1505–1518 (2022).
